# The Nature of Race in Germany: A systematic literature review of human classifications in German life sciences

**DOI:** 10.1101/2022.11.11.22282243

**Authors:** Isabelle Bartram, Laura Schnieder, Nils Ellebrecht, Florian Ruland, Tino Plümecke, Andrea zur Nieden

**Affiliations:** Institute of Sociology, University of Freiburg, Germany; Department of Biology, Chemistry, Pharmacy, Institute of Biology, Freie Universität Berlin, Germany; Leibniz-Institute of Freshwater Ecology and Inland Fisheries (IGB), Berlin, Germany

**Keywords:** Race, ethnicity, ancestry, migration background, human diversity, human classifications

## Abstract

The use of human diversity classifications like race, ethnicity, ancestry, or migration background entails a range of scientific as well as social consequences, therefore, a careful application is vital. In this article, we present results from a systematic literature review and subsequent quantitative content analysis based on 546 papers focusing on classifications applied in life sciences studies at German research institutions. Our aim is to capture a snap-shot of current classification practices applied to categorize humans across various disciplines and fields in a specific national context that remains underexposed in this regard. The review substantiates a) the results from earlier studies that point to heterogeneity, inconsistency and vagueness of human classifications used in the life sciences, and b) underlines the presumed specificity of the German science context, where the term “race” is comparatively little used. Our findings stress the need for German researchers to partake in the ongoing international debate on the practice of human classification in the life sciences to advance the international and interdisciplinary transferability of scientific results and, first and foremost, to avoid unintended effects such as overgeneralization, racialization, and stigmatization.

## Introduction

In life science research on humans, it is common to divide human diversity using categories that carry more than mere systematizing meanings. The use of terms like race, ethnicity, ancestry, or migration background can therefore result in a range of scientific as well as social consequences beyond the mere designatory function. Firstly, human categorizations such as race and ethnicity have often been criticized for not adequately reflecting biological diversity (Marks 1995; Lewontin 1972; Livingstone and Dobzhansky 1962; Roberts 2011; M’charek 2021). Secondly, classifications such as race, ethnicity, etc. inextricably contain social meanings and socio-cultural conditions of origin (Bourdieu 2018; Bowker and Star ca. 2008; Brubaker 2002; Duster 2003; M’charek 2010; Fleck 1981; Weber 1978). Thirdly, most classification terms are loaded with historical injustice, which can also make their unquestioned use in nowadays research problematic.

As a result of these criticisms editors from a variety of life science journals have sought to address these issues by publishing guidelines for potential authors (see for example Flanagin et al. 2021; Palermo et al. 2021). Beyond that, the largest professional body representing medical professionals and students in the United States, the American Medical Association, stresses that race and ethnicity are social constructs and recommend that “specific racial and ethnic categories are preferred over collective terms”. Furthermore, should the method section of articles state how participant race and ethnicity was assessed (Flanagin et al. 2021). Such kind of commitment by editors are the strongest predictor of authors following recommendations, according to a study by Sankar et al. (2015). However, Sankar et al. examine only high impact journals based in US and UK. In contrast other reviews also show that the publishing of guidelines has not necessarily led to the inclusion and/or a higher quality of reporting race/ethnicity data in the past few decades (Bokor-Billmann et al. 2020; Ma et al. 2007; Moore 2020b; Maduka et al. 2021).

In our study we give an overview of classification practices at work at German-based research institutions, elaborating mainly on the particularities of the more German versus more internationally positioned research teams and analyze the differences in the use of each term in different disciplines. This reconstruction of the respective classification practices contribute to an ongoing debate on the social repercussions of human differentiations in science.

### The seemingly absence of race in Germany

The German research context is particularly interesting because of its special relationship to the term “race” and possibly racializing human classifications. “*Rasse*” is today by no means a commonly used word in the German-speaking context. In contrast to the British and especially the US usage (race), it is now used only rarely concerning people and has almost exclusively a biological meaning (Lipphardt et al. 2018; zur Nieden 2014). There are two main reasons for this limited usage of the term in current writing. Firstly, its association with race legislation and the persecution of Jews in the Third Reich; the term “*Rasse*” itself is therefore discredited, and this resulted, secondly, in political and scientific attempts from the 1980s onwards to avoid the term and to use alternative concepts for human differentiation like ethnicity or recently migration background. In addition to the life sciences, there are also extensive efforts to replace the term race from official documents and legal texts, such as the German constitution als well as the state constitutions.

However, the race term is not simply extinguished, instead the concept and racialisations through alternative terms continue to exist, as M’charek, Schramm, and Skinner (2014) have shown also for life science research in other European countries. This is reflected by the recurring pronouncements against biological race, like the recent Jena Declaration, that can be interpreted as a persistence rather than the end of this concept. This declaration was published in 2019 by leading German zoologists and human geneticists on the occasion of the annual meeting of the German Zoological Society, and its authors demanded that “[t]oday and in the future, not using the term race should be part of scientific decency” (Fischer et al. 2019); the aim was to overcome biological justifications for racial discrimination. Evidently, statements like these continue to be necessary as the complex relationship between the life sciences and race is far from over. A wide variety of studies, mostly from the social sciences, have investigated the contemporary history of continuities of race and racializations for different life science fields in Germany such as biology (Kattmann 2002; Palm 2013; Lipphardt 2012; AG gegen Rassismus in den Lebenswissenschaften 2009), medicine (Bauer 2009; zur Nieden 2014) genetics (Plümecke 2013; Lipphardt et al. 2020; Sommer 2014) and psychology (Wandert 2009). But the majority of these studies provide rather glancing or anecdotal insights into the respective discipline and the practices of human classification in the life sciences in Germany. Up to now, no comprehensive empirical assessment of the use of human classifications in the German life sciences exists.

To address this research gap, we performed a systematic overview of studies published in recent years by German research institutions. Our aim is to provide a first overview of how human classification is currently applied in the German life sciences, under which terms, with which meanings and with what kind of differences to the international and mainly US-dominated classification schemas.

## Materials and Methods

### Systematic Literature Review

We performed a systematic literature search using PubMed and Web of Science (WoS) with the aim of identifying German life sciences studies that applied race and other possibly racializing classifications to human research subjects. In terms of method, we followed the PRISMA-P 2015 guidelines for systematic reviews and meta-analyses (see Fig.1 and checklist in supplementary material 1) (Moher et al. 2009). In our preliminary explorative research we had identified “race”, “ethnicity”, “migration background” and “ancestry” or related terms as the terms most commonly used in the life sciences to represent human diversity, and thus chose these words as search terms. The often-used term “population” was not included in our analysis because the search string led to a large number of articles that were not relevant to our research question (e.g. population-wide studies) and that could only be processed with a disproportionate amount of time.

We limited our review to primary research in English and German and excluded meta-analysis and case reports, but included studies where the authors used existing datasets to address their own research questions. We thus focused on studies where the authors themselves designed study and/or analysis protocols and potentially chose a system and terms to describe and/or differentiate research subjects.

To focus only on current studies, the search was limited to articles published between 2018 and 2020. The chosen timeframe opens up the possibility to test the potential influence of the Black Lives Matter movement in 2020 on the choice of classification terms, as indicated by interview partners from the field. This crucial question will be the focus of future investigation.The last search was performed on the 13 April 2021 to consider possible lag time from publication date to articles being indexed in scientific databases. The laborious coding process goes beyond a usual systematic review (see below) and was finished in early 2022. Using the search term “((race) OR (migra*) OR (ethnic*) OR (ancestry)) AND (German*)” we identified 3,982 records in the Pubmed database published within the selected time frame. For the WoS database the search term “race OR migra* OR ethnic* OR ancestry” was used, and the resulting search results refined to studies from Germany and from life science disciplines. 6,235 records were identified in this way. From the total of 10,217 records from both databases, 9,415 records could be excluded from further analysis because they were duplicates, the record did not meet our search criteria, or the article did not fit the scope of our review: because no human subjects were studied or artificial data was used, it did not report primary research results or addressed only single case reports, or because the subjects were not categorized or described using human diversity classifications. This strategy returned 802 relevant publications that were then read thoroughly. Another 256 publications were excluded because no full text was available via internet, library access or an email request to the authors, the studies were not from the life sciences, or any of the previously listed exclusion criteria were met (Fig. 1). The final article database contained 546 articles matching the scope of our review, 45 of them were German language (8.2%) and the rest written in English.

**Fig. 1:**
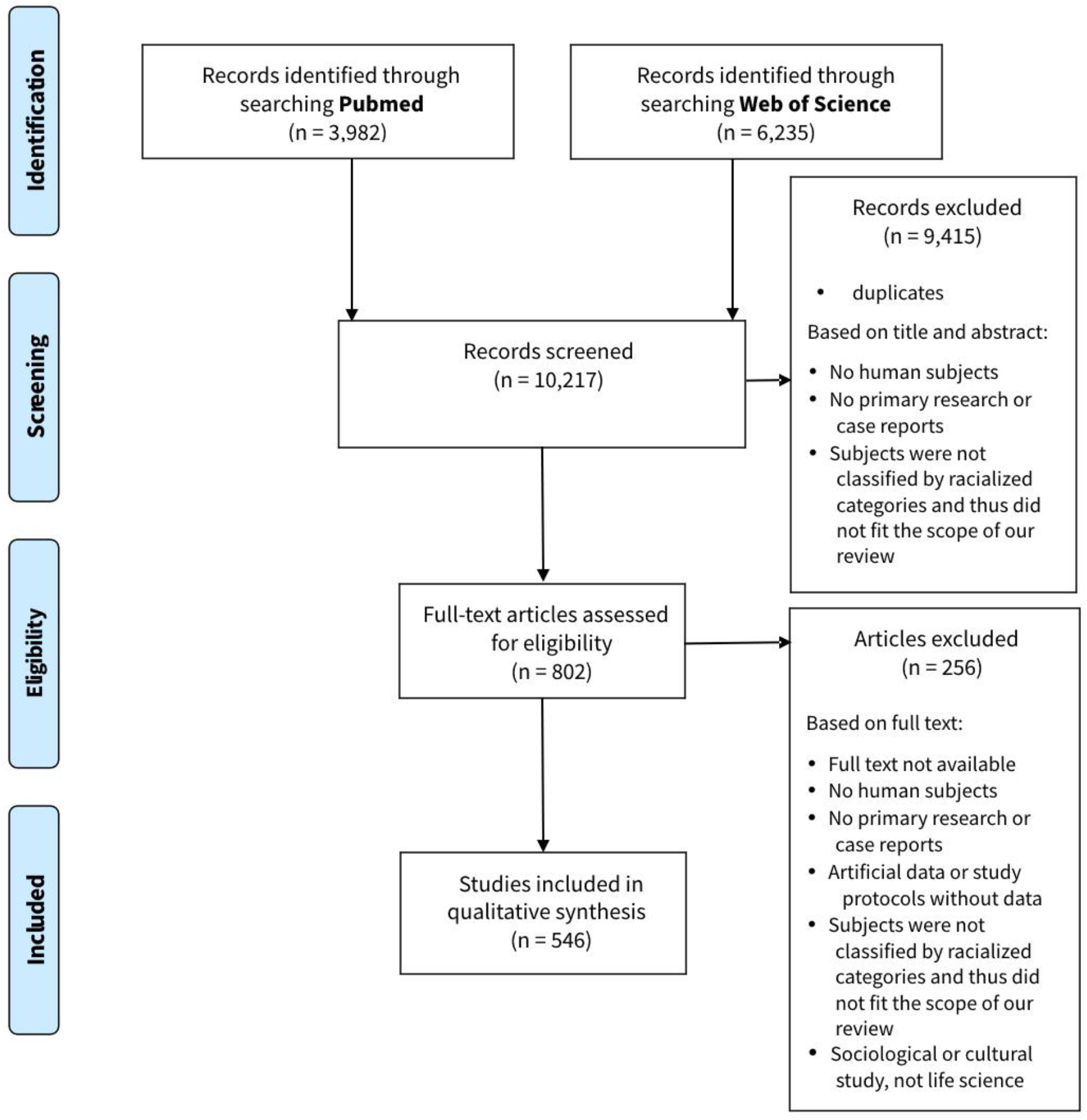
Flow chart for the selection of studies for the systematic literature review. Adapted from Moher et al. (2009)

### Quantitative analysis

To perform a quantitative content analysis (Coe and Scacco 2017), we used MAXQDA Standard 2020 20.1.0 (VERBI GmbH Berlin) software. We developed a coding system including the following categories: authors (sub-codes: all, first and last author, or first or last author affiliated to German institutes), discipline (by research institutions of first and last author), categories (which terms were used to classify or define groups of subjects), and study location (country where the samples or data were collected).

Concerning the classification terms, we focused on terms used in the methods sections to describe or stratify research subjects but also coded terms that were used by authors in the interpretation of results in the discussion section to describe the cohort they had previously analysed. Where studies did not name the location where research subjects were examined or sampled, we coded them either as “not specified” when the information was not available without guessing or as “information provided elsewhere” when the authors pointed to further research articles. Using this approach, i.e. explicitly focusing on the respective cohort, we look at the terms in context beyond a mere counting of terms.

Focusing on the methods, results and discussion sections, all articles were read thoroughly and coded by members of our research group. When uncertainties arose the publications and codes in question were discussed within our group until a consensus was achieved. The absolute and relative frequencies of coded studies were calculated for each sub-code.

We created seven generalized linear models with binomial error distribution using one of the seven most common terms (“ethnicity”, “migration”, “ancestry”, “race”, “population”, “origin” and “refugee/asylum seeker”) as respective response variable. Predictors were authors, sampling location, their interaction and the three most common disciplines (medicine, epidemiology and psychology). We performed model comparison with all possible parameter combinations and considered all predictors appearing in models with a relative model weight >/= 0.05. We built the final models using only those predictors and calculated their adjusted R^2^ values. All analyses were conducted using R (R Core Team 2020), models were created with the package “lme4” (Bates et al. 2015), model comparisons were performed using the “MuMIn” package (Bartoń) and adj. R^2^ were calculated with the package “rsq” (Zhang 2021).

## Results

### Great diversity of classification terms used in the German life sciences

With our broad search strategy, we were able to build a dataset that gives a substantial overview of the current research landscape in the respective fields. The 546 studies identified included a great variety of terms employed to stratify or describe research subjects. Within our dataset we registered 189 different classifications used. Different derivates were coded with umbrella terms, e.g. “immigrant”, “migrant”, “migration background”, and all variations and German equivalents of these terms where subsumed under the term “migration”. After merging related terms in this way, 34 different classifications could be identified within our database. Those that were applied in more than 2% of the studies analysed, and the frequency in which they were used, are listed in Table 1. The majority of studies applied several different terms to describe a cohort (54.8%), e.g. stating that people have a certain “ethnicity” or a “migration background”, often using the terms interchangeably. Of the original search terms in our systematic literature review, the classification “ethnicity” was used most often to describe the cohort studied (45.5%), followed by “migration” (33.6%), “ancestry” (17.7%) and “race” (13.9%). We identified additional terms that were used in a large proportion of studies within our database. Of these, the most common were “population” (27.8%), “origin” (14.6%), and “refugee” or “asylum seeker” (10.6%).

**Table 1:** Human classifications used in life science studies from Germany.

Great diversity of classification terms used in the German life sciences
|  |  |  |
| --- | --- | --- |
| **Search terms in literature review** | **# of studies** | **% of studies** |
| Ethnicity | 249 | 45.5 |
| Migration | 184 | 33.6 |
| Ancestry | 97 | 17.7 |
| Race | 76 | 13.9 |
| **Additional terms identified** |  |  |
| Population | 152 | 27.8 |
| Origin | 80 | 14.6 |
| Refugee / asylum seeker | 58 | 10.6 |
| (Sub)group | 36 | 6.6 |
| Descent | 30 | 5.5 |
| Nationality | 17 | 3.1 |
| Natives | 17 | 3.1 |
| Resettler / Aussiedler | 17 | 3.1 |
| Background | 13 | 2.4 |
| Language speakers | 13 | 2.4 |
| Reference group | 12 | 2.2 |
| Foreigner | 11 | 2.0 |

### German influence on classifications

We recorded whether all authors (238 studies) or the first *and* last author (119) or the first *or* last author (189) were affiliated with a German research institution. The terms used varied noticeably with the degree of ‘German-basedness’ of the author teams (Fig. 2) as well as where the study was conducted (Fig. 3). Most notably, the term “race” was not very commonly applied in studies by all-German author teams or teams with a German first and last author (4.6% and 9.2%), but used in nearly 30% of studies published by only a German first or last author. This divide was also noticeable for the terms “ethnicity” and “ancestry”. As described above, “ethnicity” was overall a more popular term than “race” within our database, and was often used alongside other terms. But while it was used in 28.6% of studies of all-German author teams, around 60% of studies by author teams less affiliated to German research institutions used it. “Ancestry” was used in only 6.3% of studies by all-German author teams, while research subjects were classified using this term in 31.1% and 23.8% of studies by a German first and last author or German first or last author. In contrast, the majority of all-German author teams used the term “migration” (57.1%) while this term was much less often used in studies by ‘less German’ author teams (22.7% and 11.1%) within our database. All above descriptive results are backed up by our quantitative analyses, as the author institution was a significant predictor in our models of the use of “ethnicity”, “race”, “ancestry” and “migration” (see supplementary material 2).

**Fig. 2:**
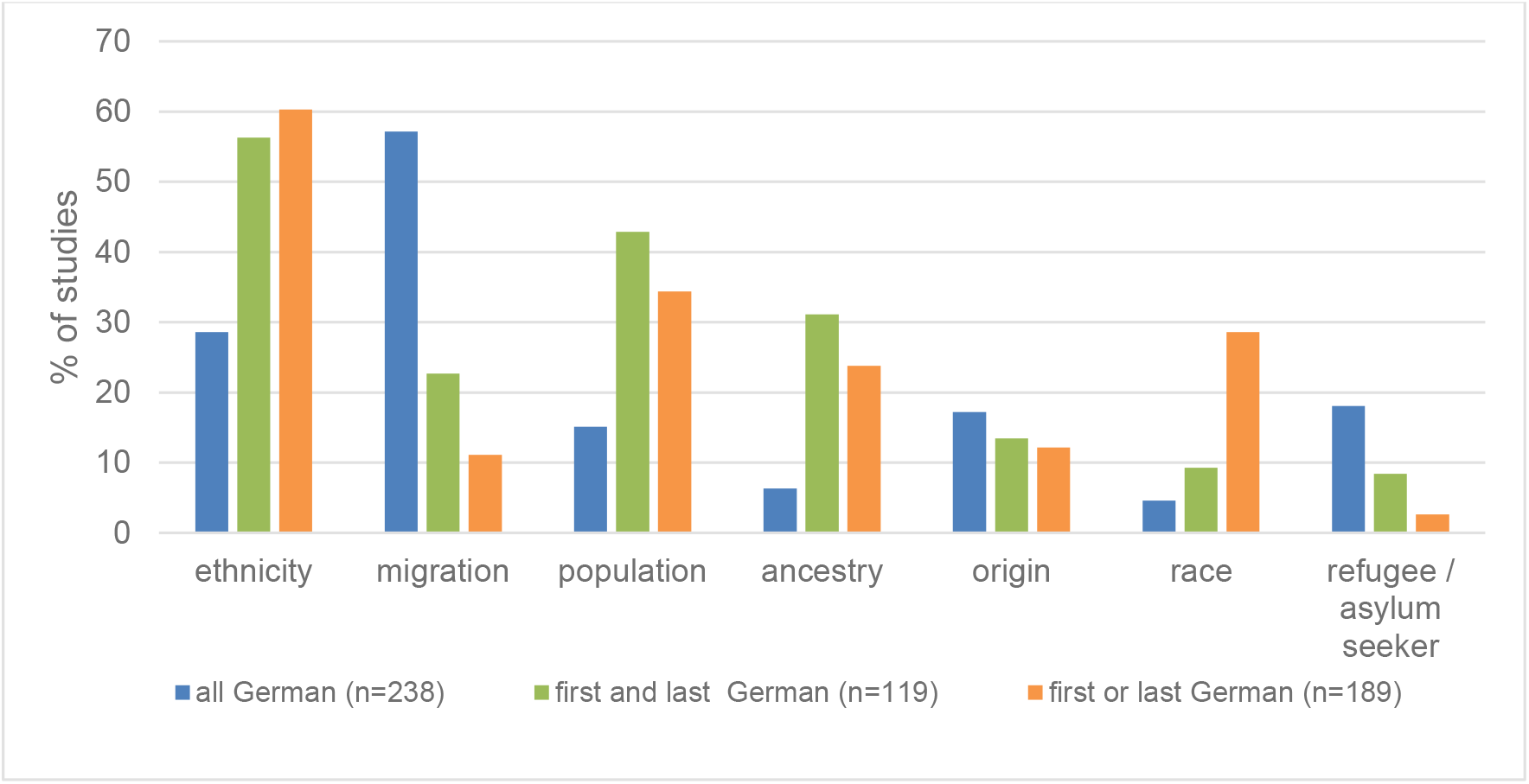
Frequency of classification terms used in relation to affiliation of author teams to German research institutions.

**Fig. 3:**
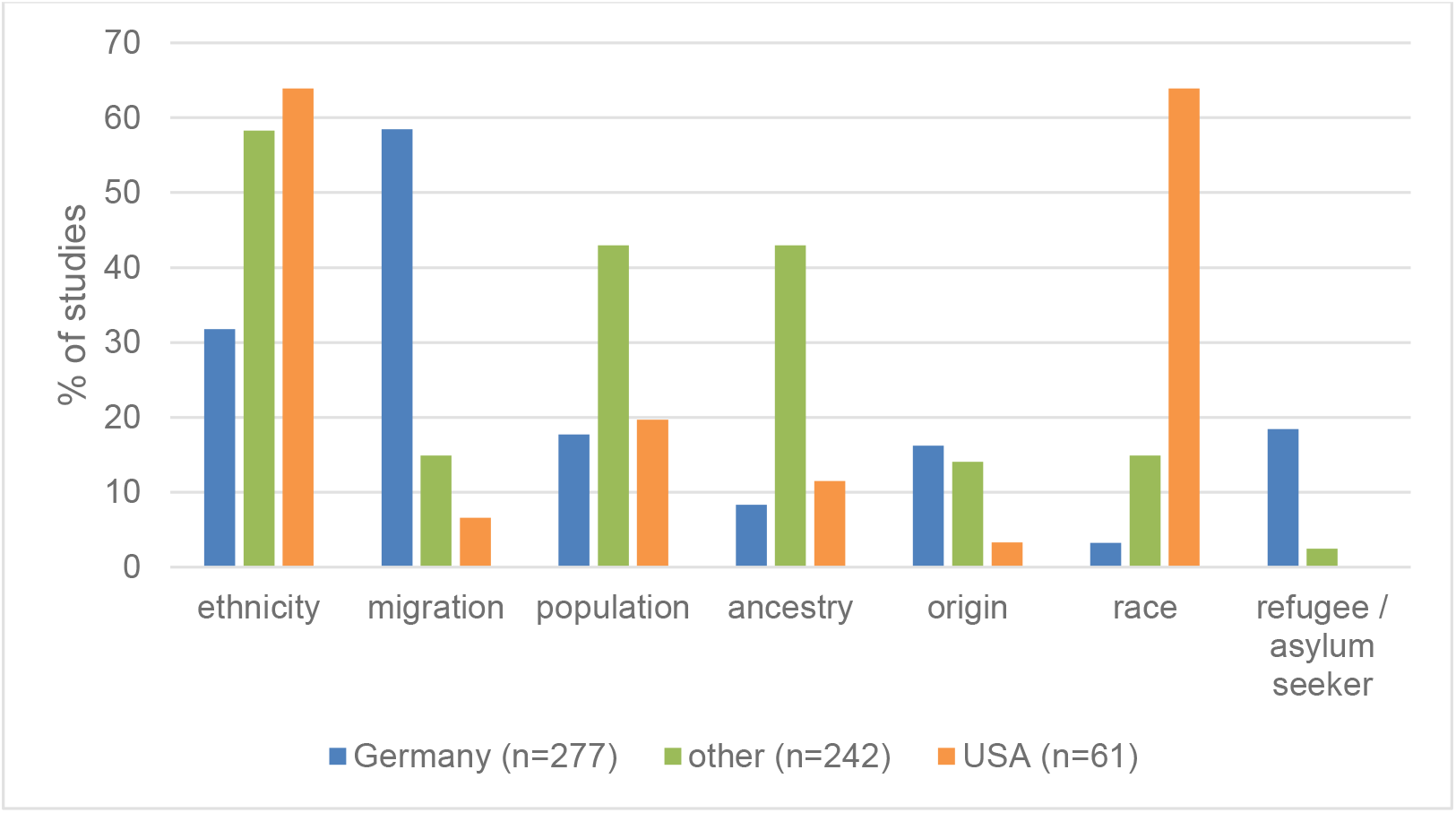
Frequency of classification terms used in relation to sampling location. Some studies examined research subjects in several countries, thus there is an overlap between articles categorized as “German”, “USA” and “other”.

The articles analysed human samples collected in 62 different countries. Most of them studied research subjects in Germany or the USA (50.6% and 11.2% of articles). Interestingly, in 35 studies (6.4%), the authors did not make sufficiently clear in which country their research subjects were examined or sampled, either by using extremely broad terms like “Europe” or “Africa” or by not mentioning a location at all.

While study authors who recruited research subjects in Germany most often described and stratified them as “migrants” or people with “migration background” (58.5%), in studies with sampling in the USA only 6.6% of studies used such a category. In contrast, 63.9% of datasets collected in the USA used the term “race” while only 3.3% of studies performed in Germany used “race” to describe research subjects (Fig. 3). Our quantitative analyses confirm this assessment as the sampling location was a significant predictor of the use of “race” (see supplementary material 2), but also for all other terms. The exceptions are population and ancestry, where the use is similar for samples from Germany or the USA, but more common if the sampling location was “other”.

### Different uses of classifications between disciplines

According to our dataset, research from different life science disciplines applies human classifications in varying frequencies to describe and differentiate research subjects (Fig. 4). Summarizing medical disciplines, we coded 42 different life science disciplines by the first and last author’s institutional or departmental affiliation. Most of the articles analysed were written by authors from the field of medicine (74.0%), followed by epidemiology (24.7%) and psychology (14.5%), with many of them published by scientists from more than one discipline (44.9%). While in the field of medicine nearly half of the author teams sorted their research subjects by their “ethnicity” (48.3%), in the epidemiology and psychology papers analysed “migration” was the most frequently used term (58.5 and 44.3%). However, neither of these terms was used frequently in the field of archaeogenetics, where “ancestry” or “population” were most often applied when grouping human research subjects (90% and 80). Our quantitative analyses confirm these observations (see supplementary material 2),

**Fig. 4:**
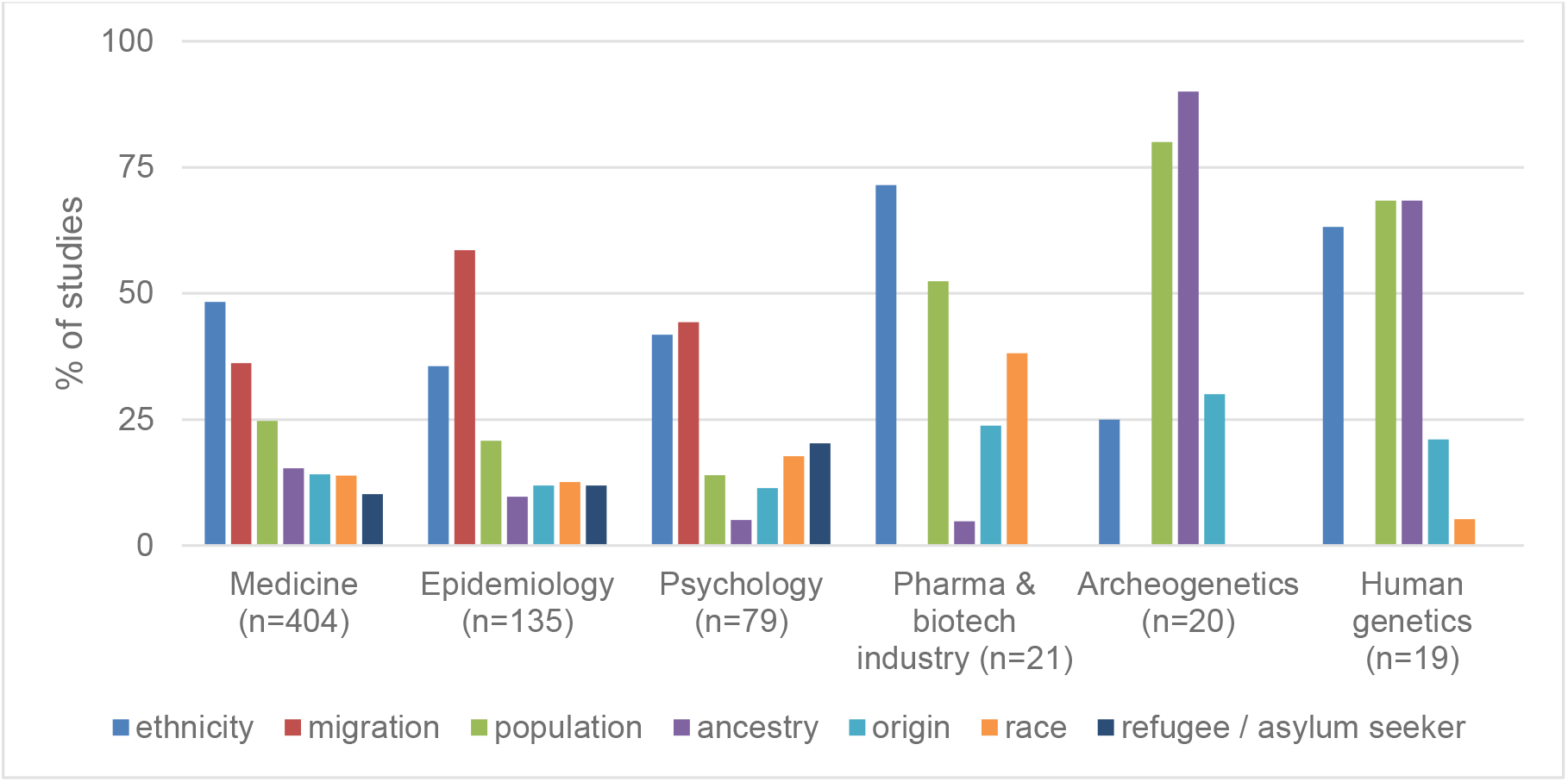
Frequency of classification terms used in different research areas. Many of the analysed studies were assigned to several disciplines, thus there is an overlap between research areas.

## Discussion

Our quantitative analysis of 546 studies published between 2018 and 2020 by authors affiliated to German institutions confirms results from smaller or more selective/specialized meta studies interested in both the wide range and the vagueness, underdetermination and inadequacy of human classifications used in the life sciences. On the other hand, our results substantiate the presumed specificity of the German science context, where the use of the term “race” is relatively low.

### Variety and inconsistencies

In their literature review, Zhang and Finckelstein examined the different racial and ethnic categories used in pharmacogenetic research (Zhang and Finkelstein 2019). They concluded that there is a high degree of heterogeneity in the categories that are used to investigate the distributions of certain genotypes. In the field of epidemiology, Bokor-Billmann et al. analysed articles in top-ranking journals for their use of ethnicity and race and identified 81 different methods authors used to classify race or ethnicity (Bokor-Billmann et al. 2020). A recent review of studies in the field of ophthalmology concluded that “the categories used were heterogeneous and often inconsistent” (Moore 2020a). Our analysis confirms these previous observations: research subjects were classified by 189 different terms, and even after we had grouped them 34 different classifications remained. Interestingly, many authors in our database used several terms next to each other (see supplementary material 2), sometimes even as proxy or synonyms. For example, Monsees et al. studied the prevalence of dementia in people with “migration background”, referencing the official definition of the German census (a person’s parents or themself born with a different nationality). But in the caption of a table summarizing the results “*Ethnien*” is used and these ethnicities are then differentiated by “*Nationalität*”, nationality (Monsees et al. 2019). While a comprehensive analysis of *how* the studies in our database defined and applied these classifications is still in progress, this finding points to a lack of awareness that terms like ethnicity, migration background and race are based on different social and academic concepts and notions.

One factor responsible for the observed divergence between studies is that while English is the standard science language, different national and cultural contexts influence classifications. For example, Zhang and Finckelstein noted a high degree of heterogeneity in the number of ethnic categories used in different countries to classify research subjects as “Asian” or “White” (Zhang and Finkelstein 2019). As anticipated, we observed a noticeable association between the degree of ‘German-basedness’ of author teams and which terms they used.

### No race in German science?

At first glance our analysis may give the impression that the German research landscape is post-racial because, firstly, the German term “*Rasse*” appears in none of the 45 research publications written in German and, secondly, the term “race” is rarely used by author teams where all members are affiliated to German research institutions compared to more international research teams (4.6 vs. 28.6%). Thirdly, race was also very rarely applied to research subjects that were sampled or examined in Germany, compared to other study locations.

However, apart from a formal application of race in the method section to classify the research subjects in the respective studies, we often observed an additional, less precise use of the term, also in the context of the cohort investigated: race is still used by many authors in the discussion section of publications when the author’s own results are compared with the findings of other studies. Kridin et al., for example, investigated specific health differences between people with Jewish and Arab ethnicity in Israel. In the discussion section the authors contrasted their results for “patients of Jewish ancestry” with a study on people of “African race”, implying that the differentiations of these two studies are comparable (Kridin et al. 2020). This kind of switching and equating of human classifications in the discussion of results is not currently covered by the usual journal guidelines. For example, the recommendations of the International Committee of Journal Editors (International Comittee of Medical Journal Editors 2021) clearly only refer to the respective study participants when recommending defining how race or ethnicity should be determined and the use of precise language.

Instead of race and ethnicity, author teams affiliated to German research institutions often use the term “migration” (including migrant, immigrant, migrant background etc.) to describe and stratify their research subjects. Further investigation would be needed to establish whether “migration” is simply used as a synonym to replace socially controversial terms, or the use of race and ethnicity is low because German scientists seldomly see any scientific or ethical benefit in applying it. Preliminary results of our analysis for the field of epidemiology show that, on the one hand, the choice of categories also relates to which categories of differentiation are used in the political sphere. Germany, unlike the US, has no ethnic census categories but recently introduced the variable “migration background” for the micro census to record first, second or third generation immigrants to Germany that may have German citizenship and are no longer trackable by the former category “*Ausländer*” (foreigner) (zur Nieden and Bartram 20.08.2020; zur Nieden 2014). Subsequently, German epidemiological studies have begun to apply this category if samples are collected in Germany instead of race or ethnicity, arguing e.g. that Germany is “an immigration country without post-colonial migration and without numerically relevant autochthonous ethnic minorities” (Schenk 2007). On the other hand migration background is often used interchangeably with ethnicity, maybe to ensure compatibility with the international discussion (zur Nieden and Bartram 20.08.2020).

Finally, it can be noted that the terms “refugee” and “origin” are used more frequently the more German authors are involved in the study. The use of the former possibly reflects the research interest in the situation of refugees after the increased social debates on immigration from 2015 on. The predilection for the English term “origin”, which is rarely used by native speakers, can in turn be understood as an expression of linguistic uncertainty. One can even suspect that the underdetermined nature of the term encourages its deliberate use.

### Disciplinary classification cultures

Our literature review yielded studies from a broad variety of fields in the life sciences, including clinical studies of pharmaceutical/biotech companies. Nearly half of them were published by authors from different research fields, speaking to a relatively high degree of interdisciplinarity of the German research landscape on human diversity. However, nearly 3/4 of studies were affiliated to medical departments. Whether this dominance of medical studies is due to a general dominance of medicine in German life science research (due to research funding allocation and publication cultures), or whether medical studies are more likely to apply human classifications, is unclear. Due to the low number of studies from disciplines besides medicine, epidemiology, and psychology our results should be taken with a grain of salt. Nonetheless, while these three disciplines show a rather similar frequency distribution of the classification terms ethnicity, ancestry, race and migration; we observed a different practice in other disciplines. Studies from archaeogenetics and human genetics, for example, mainly refer to “ancestry” to describe or divide their research subjects. Here, “ancestry” is usually genetically determined, and interestingly is sometimes even used to verify the self-reported ethnicity of research subjects (Degenhardt et al. 2019). This is in accordance with observations by Fujimura and Rajagopalan, who studied population geneticists in the US [29]. Most studies in these fields, especially when they are investigating human genetic history, are conceived by large international author teams. Thus, the unpopularity of the term race can be attributed to disciplinary culture and not to German scientific culture (Byeon et al. 2021; Popejoy et al. 2020).

In contrast, one third of the 17 clinical studies by pharmaceutical companies within our database used the term race (only 13.9% on average in the whole database). This is most certainly driven by US standards, as the US Food and Drug Administration demands reporting of race/ethnicity for drug approvals. How this classification is then applied to a German study population and to what effect remains to be investigated.

### Limitations

Certainly, important classification terms like population and origin are underrepresented in our review sample, as the terms generated a plethora of non-specific search results that we did not want to include in our review or could not meaningfully evaluate with the resources available to us. In addition, the decisions we took in determining what constitutes the “German life sciences” may have had an effect on the inclusion or exclusion of some research papers. Publication language is not a useful indicator as, apart from epidemiological studies, today relevant scientific results from German research institutions are published almost exclusively in English-language journals (Baethge 2008). Additionally, research teams at German institutions are frequently very international, e.g. in 2017 nearly half of the scientists employed at the Max Planck Society did not have German nationality (Deutscher Akademischer Austauschdienst and Deutsches Zentrum für Hochschul-und Wissenschaftsforschung 2020). In addition, scientific résumés demand work abroad during the postdoctoral phase of German scientists, possibly with a lasting impact on their research activity. Thus, affiliation to a German institution might be an imprecise surrogate marker in studying historical and cultural differences along national and language lines. Similarly, it was sometimes difficult to determine the discipline of a given study without expertise in some of the specific research topics. Thus, as this article is published in a science journal by an interdisciplinary author team, mainly affiliated to a sociological institute, we are aware that disciplinary subtleties might have been overlooked by our approach. However, we are confident that the search strategy of our literature review captured a broad overview of life science studies affiliated to German research institutes regarding our research question.

## Conclusion

Our quantitative content analysis of research literature illustrates that current German life sciences are prone to the same heterogenous use of human classifications that was found in the international studies cited. Additionally, we identified a unique relationship with the term “race” that was used less the more German-based an author team was. The often interchangeable use of terms like race, ethnicity, ancestry, migration etc. we registered within the same studies is an indication of a confusion and underdetermination of concepts that can lead to overgeneralization and stigmatization. Regardless of whether human classifications are included in research to measure and combat health inequalities, trace human history or identify population-specific therapeutic targets, terms like race, ethnicity and migration background cannot be separated from their social meanings. Furthermore, imprecise and vague classifications also impede the national and international transferability of scientific results, thereby diminishing even the intended impact of the research in question.

We are very well aware that the negotiation and decision-making processes inherent in every step of our analysis from outline to coding practices to the formulation of results gives this review a very specific angle. And of course, some of the decisions made (e.g. how to demarcate which publications count in as *German* research) are as imperfect and as much subject to negotiation as the classifications analysed themselves. Still, we are convinced that this overview and the aspects we highlighted serve as a worthwhile basis to get involved into a trans- and international debate about the potentials and pitfalls of classification practices in the current life sciences.

## Supporting information

Supplementary Material 1: PRISMA 2009 checklist.

Supplementary Material 2: Model selection

## Data Availability

All data used in the present study are available upon reasonable request to the authors.

## Acknowledgments

We thank Isabel Schön, Felix Fink and Charlotte Schulze-Marmeling for their assistance in compiling and coding the articles in our database.

This study was supported by a grant by the German Federal Ministry of Education and Research (BMBF), funding number 01GP1790.

## Supporting information

**Supplementary Material 1: PRISMA 2009 checklist**.

From: Moher D, Liberati A, Tetzlaff J, Altman DG, The PRISMA Group (2009). Preferred reporting items for systematic reviews and meta-analyses: The PRISMA statement. PLoS Med 6(7): e1000097. doi:10.1371/journal.pmed1000097 https://doi.org/10.1371/journal.pone.0225898.s002

**Supplementary Material 2: Model selection**

