## Supplementary Material 2: Model selection for "The Nature of Race in Germany: A systematic literature review of human classifications in German life sciences"

### Model Selection

sl = sampling location

au = author location

med = medicine

epi = epidemiology

psy = psychology

#### Model 1: ethnicity

Full model:

ethnicity ~ samplingLocation + authorLocation + samplingLocation:authorLocation + medicine + epidemiology + psychology [*model S1a*]

Table S1: Model selection output of model S1. Models sorted by delta AIC. All predictors appearing in all models with an AIC weight of >/= 0.05 are highlighted in **bold**.

| Model (predictors) | delta-AIC | AIC weight |
|---|---|---|
| **sl + au** + sl:au + med - epi | 0.00 | 0.16 |
| **sl + au** + sl:au + med | 0.57 | 0.121 |
| **sl + au** + med - epi | 0.81 | 0.107 |
| **sl + au** + sl:au + med - epi + psy | 1.40 | 0.08 |
| **sl + au** + sl:au + med + psy | 1.41 | 0.079 |
| **sl + au** + sl:au - epi | 1.47 | 0.077 |
| **sl + au** +med | 1.72 | 0.068 |
| **sl + au** + med - epi + psy | 1.95 | 0.06 |
| **sl + au** + med + psy | 2.21 | 0.053 |
| **sl + au** + sl:au | 2.26 | 0.052 |

Final model:

ethnicity ~ samplingLocation + authorLocation [*model S1b*]

adj. $R^2$=0.1

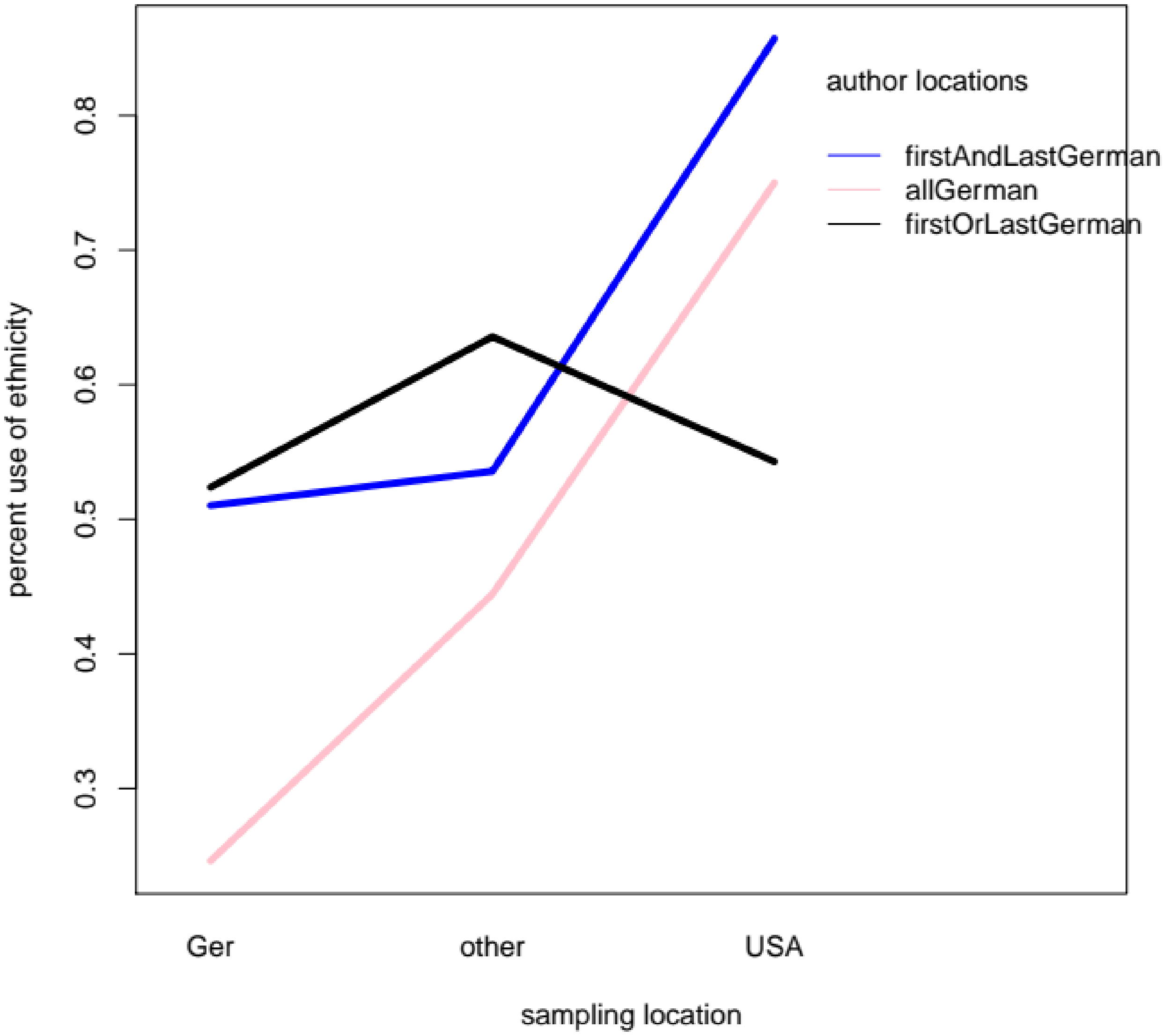


*Fig. S1: Interaction plot of the percent of the term ethnicity used across sampling locations and author locations.*

### Model 2: migration

Full model:

migration ~ samplingLocation + authorLocation + samplingLocation:authorLocation + medicine + epidemiology + psychology [*model S2a*]

| **Model (predictors)** | **delta-AIC** | **AIC weight** |
|---|---|---|
| **sl + au** + med **+ epi** + psy | 0 | 0.372 |
| **sl + au** + med **+ epi** | 1.13 | 0.212 |
| **sl + au** + sl:au + med **+ epi** + psy | 1.74 | 0.156 |
| **sl + au** + sl:au + med **+ epi** | 2.82 | 0.091 |
| **sl + au + epi** | 3.51 | 0.064 |

Final model:

migration ~ samplingLocation + authorLocation + epidemiology [*model S2b*]

adj. $R^2$ = 0.35

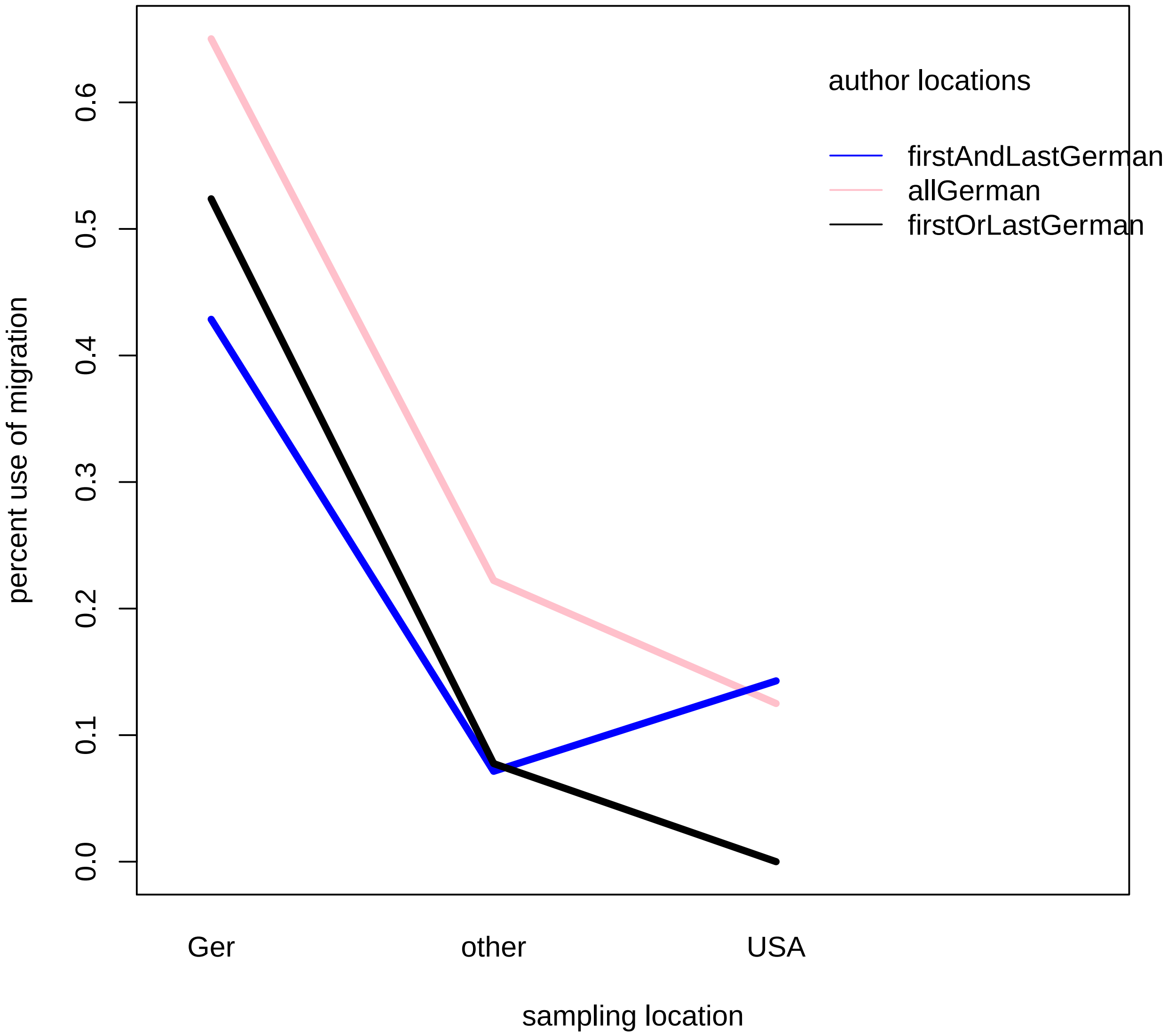


*Fig. S2: Interaction plot of the percent of the term migration used across sampling locations and author locations.*

**Model 3: ancestry-1.04500 + -0.4041 -0.6990 -1.0380 + 8 -282.190 580.6 0.00**

30 -1.22400 + -0.6583 -0.9428 + 7 -283.428 581.1 0.42

Full model:

ancestry ~ samplingLocation + authorLocation + samplingLocation:authorLocation + medicine + epidemiology + psychology [*model S3a*]

| Model (predictors) | delta-AIC | AIC weight |
|---|---|---|
| **sl + au** - med - epi **- psy** | 0 | 0.489 |
| **sl + au** - epi **- psy** | 2.28 | 0.156 |
| **sl + au** - med **- psy** | 2.31 | 0.154 |
| **sl + au - psy** | 4.48 | 0.052 |

Final model:

ancestry ~ samplingLocation + authorLocation + psychology [*model S3b*]

adj. $R^2 = 0.16$

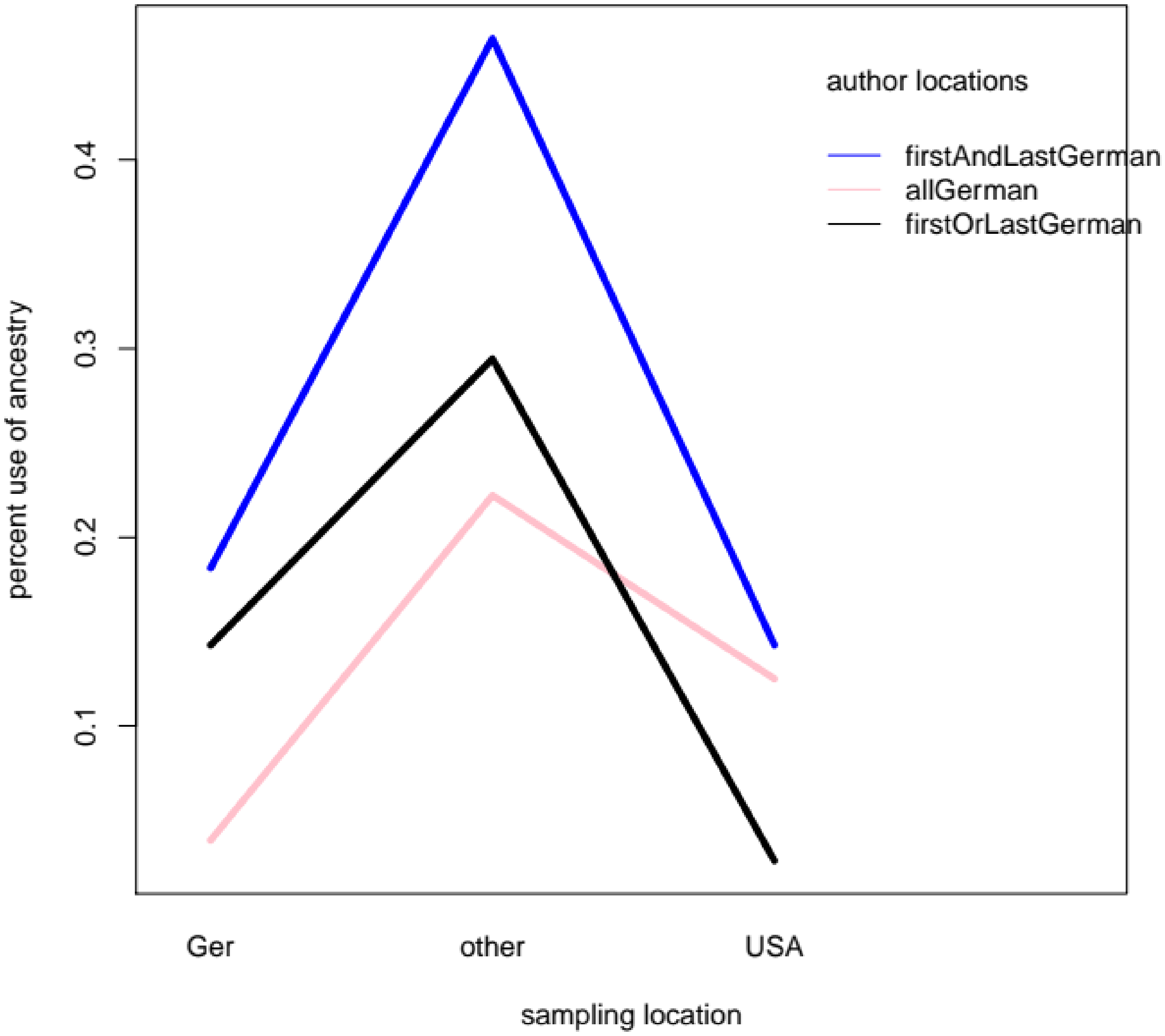


*Fig. S3: Interaction plot of the percent of the term ancestry used across sampling locations and author locations.*

### Model 4: race

Full model:

race ~ samplingLocation + authorLocation + samplingLocation:authorLocation + medicine + epidemiology + psychology [*model S4a*]

| Model (predictors) | delta-AIC | AIC weight |
|---|---|---|
| **sl + au + psy** | 0.00 | 0.23 |
| **sl + au** + sl:au **+ psy** | 0.89 | 0.149 |
| **sl + au** - med **+ psy** | 1.02 | 0.14 |
| **sl + au** + sl:au - med **+ psy** | 1.93 | 0.088 |
| **sl + au** - epi **+ psy** | 2.04 | 0.084 |
| **sl + au** + sl:au - epi **+ psy** | 2.95 | 0.053 |
| **sl + au** - med - epi **+ psy** | 4.06 | 0.05 |

Final model:

race ~ samplingLocation + authorLocation + psychology [*model S4b*]

adj. $R^2 = 0.32$

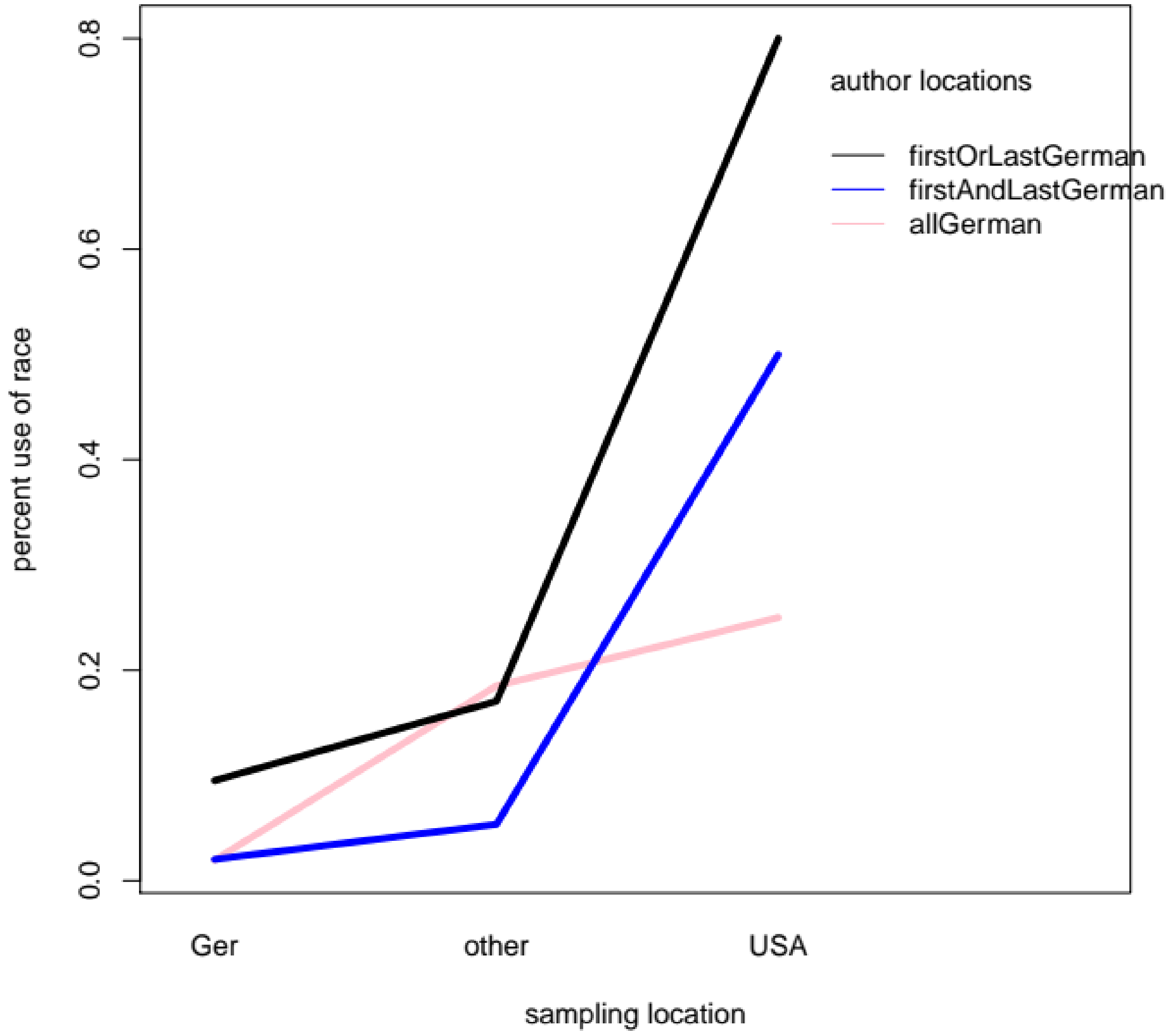


*Fig. S4: Interaction plot of the percent of the term race used across sampling locations and author locations.*

### Model 5: population

Full model:

population ~ samplingLocation + authorLocation + samplingLocation:authorLocation + medicine + epidemiology + psychology [*model S5a*]

| Model (predictors) | delta-AIC | AIC weight |
|---|---|---|
| **sl + au - med** - epi **- psy** | 0 | 0.461 |
| **sl + au - med - psy** | 0.42 | 0.375 |

Final model:

population ~ samplingLocation + authorLocation + medicine + psychology [*model S5b*]

adj. $R^2 = 0.13$

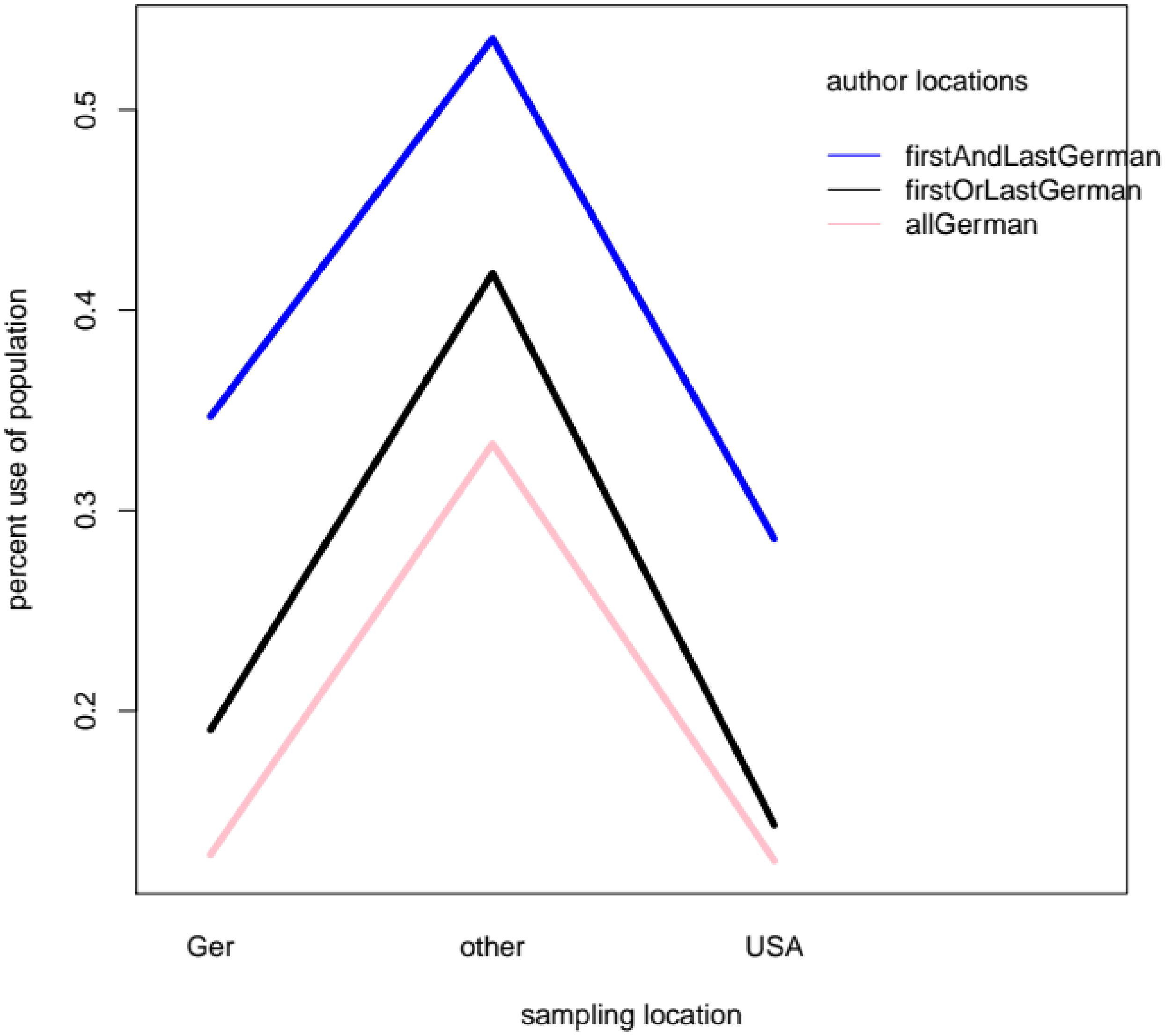


*Fig. S5: Interaction plot of the percent of the term population used across sampling locations and author locations.*

### Model 6: origin

Full model:

origin ~ samplingLocation + authorLocation + samplingLocation:authorLocation + medicine + epidemiology + psychology [*model S6a*]

| **Model (predictors)** | **delta-AIC** | **AIC weight** |
|---|---|---|
| **sl** | 0 | 0.196 |
| **sl** - epi | 0.86 | 0.128 |
| **sl** - psy | 1.08 | 0.114 |
| **sl** - epi - psy | 1.51 | 0.092 |
| **sl** - med | 2 | 0.072 |
| **sl** +au | 2.51 | 0.056 |

Final model:

origin ~ sampling Location [*model S6b*]

adj. $R^2 = 0.01$

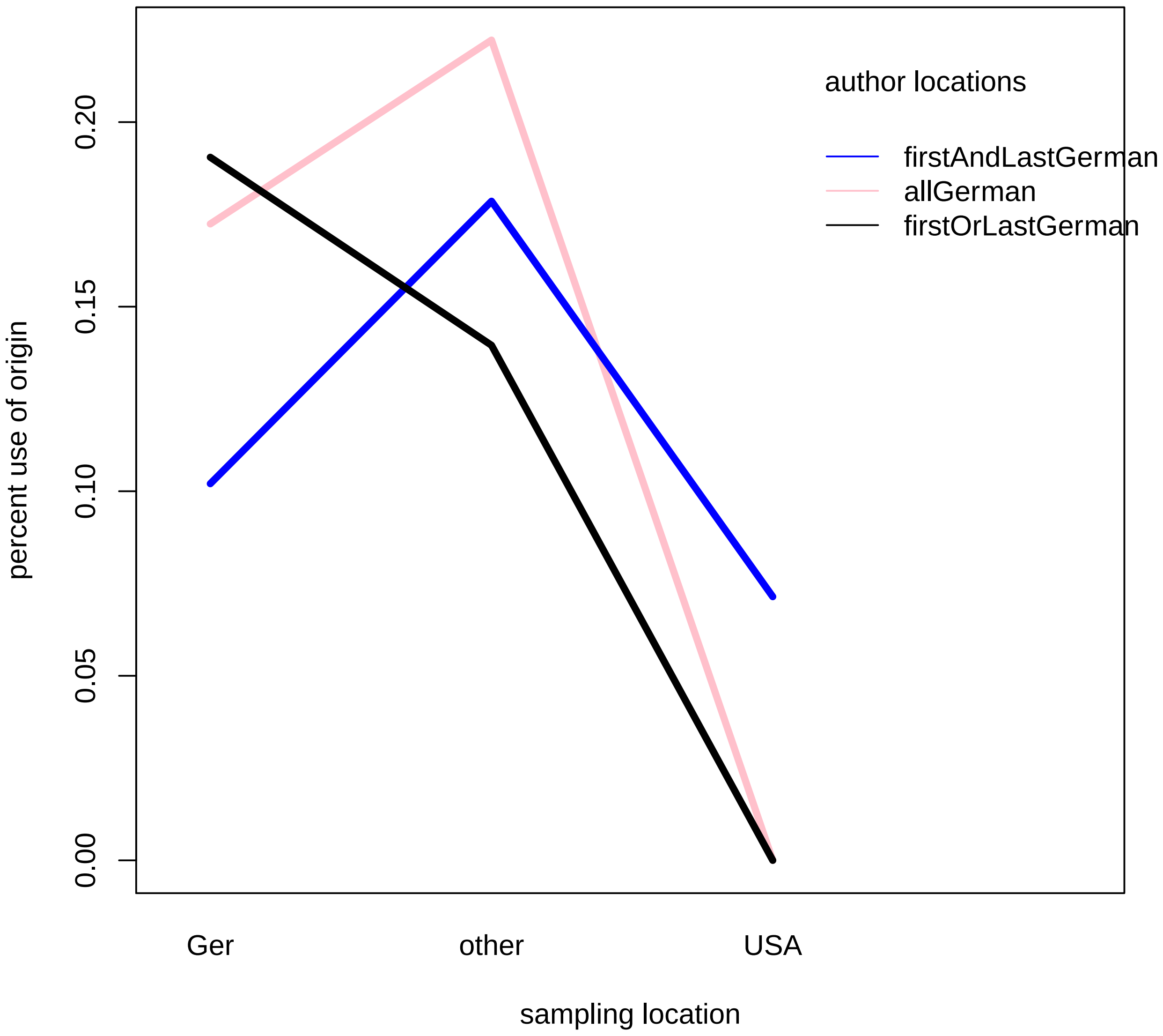


*Fig. S6: Interaction plot of the percent of the term origin used across sampling locations and author locations.*

### Model 7: refugee / asylum seeker

Full model:

refugeeAsylumSeeker ~

samplingLocation + authorLocation + samplingLocation:authorLocation +

medicine + epidemiology + psychology [*model S7a*]

| Model (predictors) | delta-AIC | AIC weight |
|---|---|---|
| **sl**+ psy | 0 | 0.245 |
| **sl** + au + psy | 1.41 | 0.121 |
| **sl** | 1.87 | 0.096 |
| **sl** - med + psy | 2.03 | 0.089 |
| **sl** + epi + psy | 2.04 | 0.088 |
| **sl** +au | 2.8 | 0.06 |

Final model:

refugeeAsylumSeeker ~ samplingLocation [*model S7b*]

adj. $R^2 = 0.07$

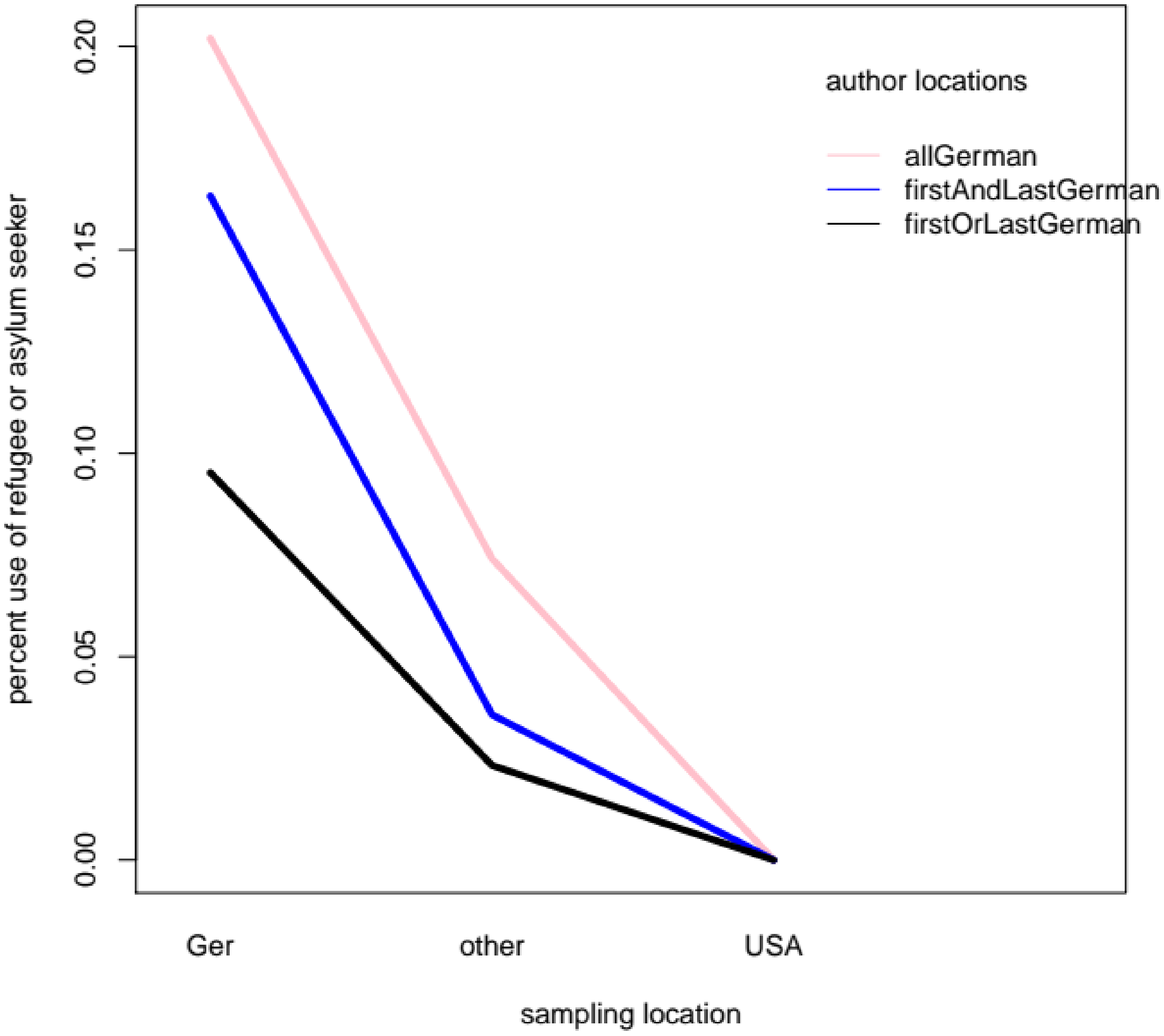


*Fig. S7: Interaction plot of the percent of the terms refugee / asylum seeker used across sampling locations and author locations.*

### Distribution across disciplines

Most studies were part of the disciplines medicine, epidemiology and psychology. Other disciplines, like industry or archaeogenetics, did not have a balanced enough distribution across sampling locations or author locations. Therefore, only medicine, epidemiology and psychology were analyzed in depth.

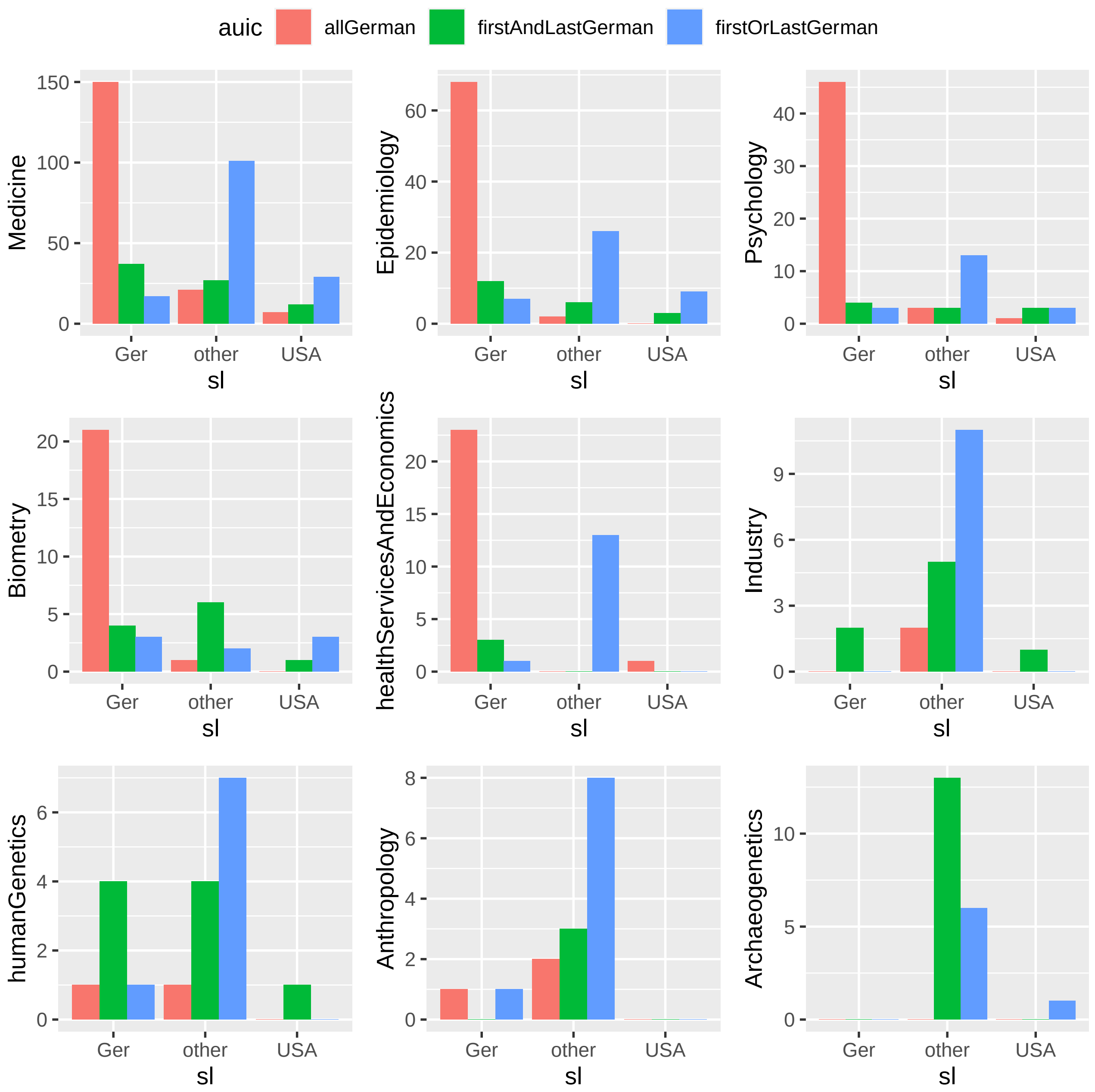


*Fig. S8: Distribution of studies across disciplines, author locations and sampling locations.*

### Author networks

Almost all authors appeared only once or very few times either as first- or last authors. This leads us to assume there is little bias in personal styles of word use by authors, although author networks can still play a role.

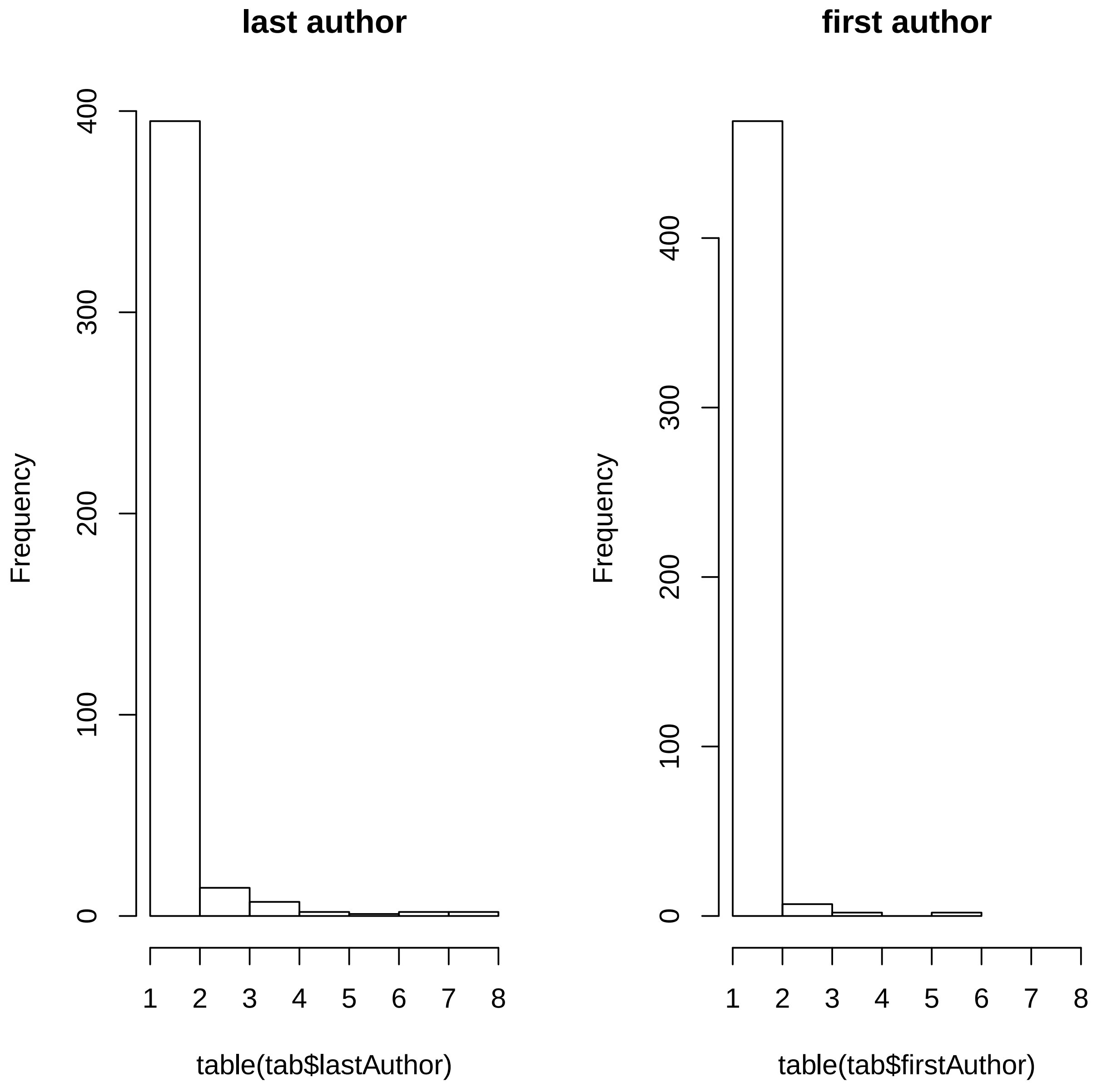


*Fig. S9: Number of appearances of authors as either last- or first author in our dataset.*
